# Deep learning predicts HRD and platinum response from histology slides in breast and ovarian cancer

**DOI:** 10.1101/2023.02.23.23285869

**Authors:** Erik N. Bergstrom, Ammal Abbasi, Marcos Díaz-Gay, Loïck Galland, Scott M. Lippman, Sylvain Ladoire, Ludmil B. Alexandrov

## Abstract

Breast and ovarian cancers harboring homologous recombination deficiencies (HRD) can benefit from platinum-based chemotherapies and PARP inhibitors. Standard diagnostic tests for detecting HRD utilize molecular profiling, which is not universally available especially for medically underserved populations. Here, we trained a deep learning approach for predicting genomically derived HRD scores from routinely sampled hematoxylin and eosin (H&E)-stained histopathological slides. For breast cancer, the approach was externally validated on three independent cohorts and allowed predicting patients’ response to platinum treatment. Using transfer learning, we demonstrated the method’s clinical applicability to H&E-images from high-grade ovarian tumors. Importantly, our deep learning approach outperformed existing genomic HRD biomarkers in predicting response to platinum-based therapies across multiple cohorts, providing a complementary approach for detecting HRD in patients across diverse socioeconomic groups.

**One-Sentence Summary:** A deep learning approach outperforms molecular tests in predicting platinum response of HRD cancers from histological slides.

## MAIN TEXT

Precision oncology aims to personalize cancer therapy by first identifying and, subsequently, targeting molecular defects in tumors within each individual (*1*). Many cancers harbor failures of specific DNA repair pathways and utilizing synthetic lethal relationships amongst peripheral pathways has proven an effective treatment approach (*2*). Previous mechanistic studies and clinical trials have shown that breast and ovarian cancers harboring homologous recombination DNA repair deficiency (HRD) are highly sensitive to platinum salts and poly (ADP-ribose) polymerase (PARP) inhibitors (*3*). Historically, HRD has been associated with germline mutations in specific genes leading to an increased cancer risk with the most notable susceptibility genes being *BRCA1* (*4*) and *BRCA2* (*5*). In addition to germline variants, somatic mutations and epigenetic dysregulation can also lead to HRD (*6*). Importantly, HRD cancers exhibit characteristic patterns of somatic mutations (*6-10*) and gene expression (*11, 12*), and these patterns have been leveraged as predictive biomarkers for targeted response to platinum therapy and PARP inhibitors. Notably, the pattern of single-base substitution signature 3 (SBS3), part of the Catalogue of Somatic Mutations in Cancer (COSMIC) catalog of mutational signatures (*13*), was previously utilized as a clinical biomarker for detecting HRD (*7, 14*).

In the United States, the FDA has approved two HRD companion diagnostic (CDx) tests for patients with ovarian and metastatic breast cancer (*15*). Myriad myChoice® CDx and FoundationOne® CDx determine HRD by quantifying overall genomic instability in combination with *BRCA1/2* status (*16, 17*). Additionally, multiple research and CLIA-certified HRD diagnostic tests have been developed (*18*) and utilized to characterize the prevalence of HRD across different solid tumors (*19-21*). These studies have identified HRD as commonly found in multiple refractory human cancers, including triple-negative breast cancer, high-grade serous ovarian cancer, and pancreatic adenocarcinoma, as well as established the role of HRD-targeted therapies with platinum salts and PARP inhibitors (*16, 22-24*). Currently, all existing HRD diagnostic tests intrinsically rely on DNA and/or RNA profiling leading to clinical-workflow bottlenecks largely attributed to the availability of sufficient tissue samples for molecular assays as well as to time to decision making and overall cost (*25-27*). For example, the cost of an FDA-approved or a CLIA-certified HRD test is several thousand dollars (*27*) and results can take from 3 to 6 weeks (*25*). In turn, this has precluded the widespread utilization of molecular diagnostics in standard therapy and clinical trials (*1*) with a disproportionately high effect on patients from underserved populations (*15*).

While the utilization of sequencing-based diagnostics is limited, tumor biopsies are routinely processed in clinical practice for the diagnosis of solid-tumors by light-microscopic morphological review of tissue stained with hematoxylin and eosin (H&E) (*26*). Combined with recent advances in computational pathology, deep learning artificial intelligence (AI)-based models allow for both prognostic and diagnostic predictions using only digital H&E slides (*28*). Here we introduce DeepHRD, a deep learning AI platform for detecting HRD from digitalized H&E slides. We train and validate DeepHRD models on data from The Cancer Genome Atlas (TCGA) project and demonstrate their ability to detect HRD using external datasets. Importantly, using independent samples, we demonstrate that DeepHRD outperforms existing clinical genomic biomarkers in predicting response to platinum-based therapies.

## RESULTS

We implemented DeepHRD, a weakly supervised convolutional neural network architecture that uses multiple instance learning (MIL; **Fig. 1**) for predicting HRD status from digital H&E slides (*29-31*). Specifically, for training DeepHRD, a soft label is assigned to each digital whole-slide H&E image (WSI) based on an HRD score derived using sequencing or genotyping data from the same cancer sample (**Supplementary Materials**). Further, all partitioned regions of that WSI, termed, tiles, are assigned a weak label based upon the sample’s classification. It is assumed that all tiles within a negatively labeled sample are homologous recombination proficient (HRP), whereas at least one tile must exhibit an HRD phenotype within a positively labeled sample. These assumptions allow the model to be trained using only a single classification label for an entire WSI without the need for detailed manual annotations from a pathologist, which currently do not exist for characterizing HRD.

**Fig. 1.**
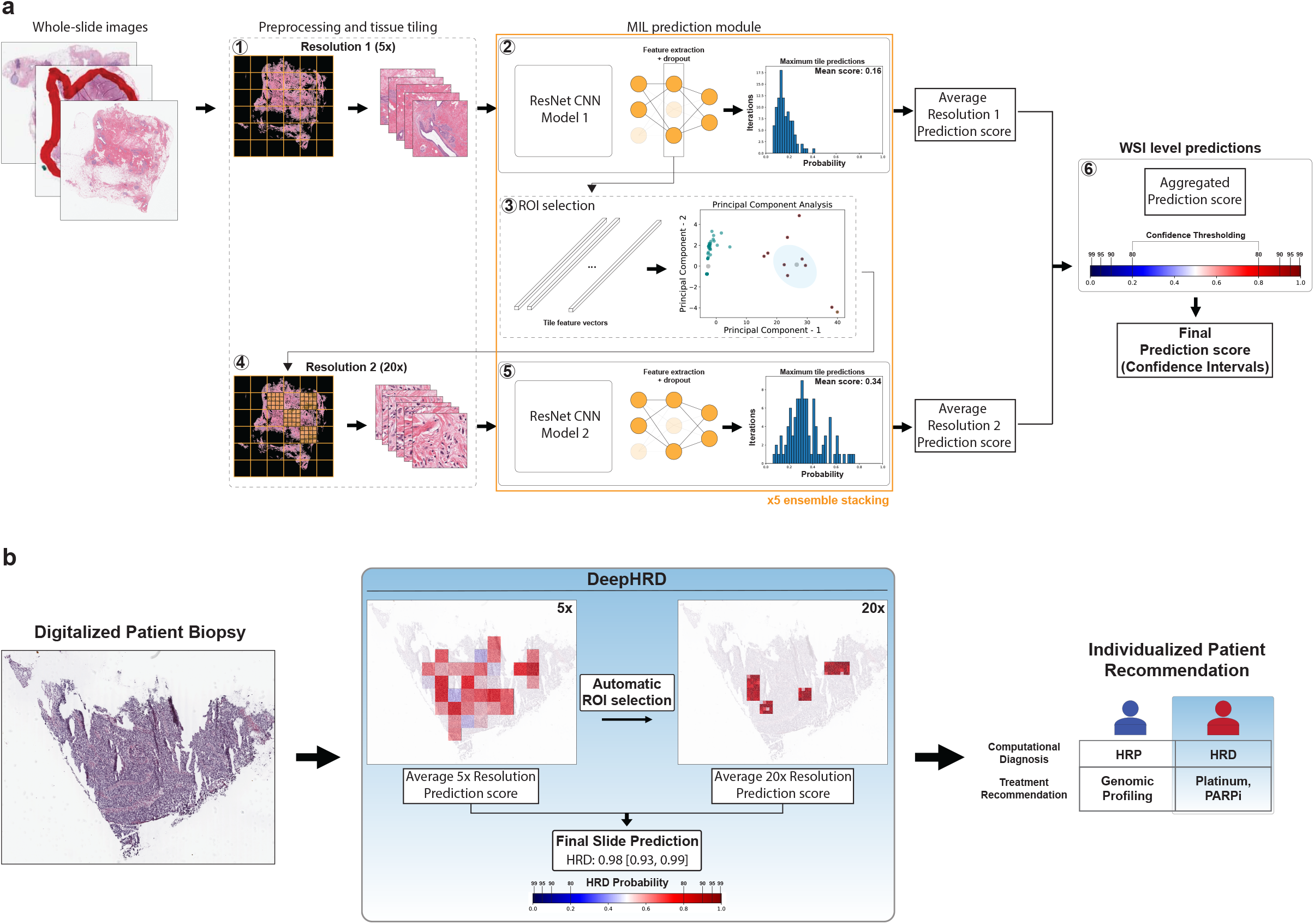
Multi-resolution convolutional neural network architecture to detect homologous recombination deficiency from histopathological tissue slides. ***a)*** Training a DeepHRD model for detecting homologous recombination deficiency (HRD) from whole-slide images (WSIs). For each WSI, a single prediction score is estimated based on the detection of HRD. Specifically, each WSI undergoes preprocessing and quality control (**1**). This module consists of tissue segmentation, filtering for non-focused tissue, and final tiling of regions that contain tissue at 5x magnification. All tiles for a single image are processed through the first multiple instance learning (MIL) ResNet18 convolutional neural network (**2**). This architecture uses the average of the top 25 predicted tile scores as the WSI predicted score. Dropout is incorporated into the fully connected layers in the feature extraction module to reduce overfitting during training. The same dropout technique is also incorporated during inference to simulate Monte Carlo dropout used to calculate confidence intervals in the final WSI prediction. The tile feature vectors from the penultimate layer of the feature extraction are used to automatically select regions of interest (ROI) from the original WSI for additional assessment (**3**). The feature vectors are reduced in dimensions using principal component analysis and a custom k-means clustering module is used to determine the optimal number of clusters per sample. The selected tiles are then resampled at a 20x magnification (**4**). These sets of tiles are used to train a second MIL-ResNet18 model (**5**) using an identical architecture to the one previously used in (**2**). The average predictions across both models are aggregated for a single WSI (**6**). The resulting distribution of scores are used to calculate confidence intervals and establish a threshold of confidence for a final prediction. ***b*)** Using a trained DeepHRD model for HRD prediction from a single whole-slide image. DeepHRD produces a final prediction score for individual patient biopsies, with a computational-based diagnosis for subsequent clinical action.

DeepHRD is based on a multi-resolution decision designed to mimic the standard diagnostic protocol used by pathologists, which performs an initial prediction on a low magnification (*i*.*e*., 5x magnification) and then automatically selects regions of interest (ROI) to perform a secondary prediction on an enhanced magnification within ROIs (*i*.*e*., 20x magnification; **Fig. 1*a***) (*30*). Once fully trained, the model generates HRD predictions directly from digital tissues slides without the need for genomic profiling (**Fig. 1*b***). Further, DeepHRD maps individual tile predictions back to the original WSI, which allows visualizing the relative importance of tissue regions for the obtained predictions (**Fig. 1*b***). The final model encompasses an ensemble of five identical architectures, with each producing multi-resolution prediction scores. The average of these scores is used to make a final prediction for each tissue slide. Importantly, DeepHRD estimates epistemic uncertainty using Bayesian dropout during inference of a tissue slide to calculate confidence intervals for the final model prediction (**Supplementary Materials**). The confidence intervals are subsequently used to provide a computational diagnostic recommendation (**Fig. 1*b***).

DeepHRD models were trained and internally validated using data from 1,008 TCGA breast cancers (*32*) with flash frozen (FF) slides and 1,055 TCGA breast cancers with formalin-fixed paraffin-embedded (FFPE) slides (**fig. S1**). All samples had whole-exome sequencing and microarray genotyping data for calculating a genomic HRD score (**fig. S1**). We trained DeepHRD breast cancer models by separating the samples with: *(i)* 70% used for training; *(ii)* 15% for adjusting training parameters; and *(iii)* 15% held-out for testing the final model (**Fig. 1*a***; **fig. S1**). Two independent models were trained, one for FF and one for FFPE tissue slides (**Supplementary Materials**). Prior to training, the number of HRD and HRP samples per breast cancer subtype were balanced to prevent learning subtype specific histological features (**fig. S1**). Each trained DeepHRD breast cancer model allows making a patient-level prediction using only a single FF or FFPE digital slide. Specifically, DeepHRD predicts whether a breast cancer is HRD or HRP, and it overlays an HRD probability mask to the digital slide, thus, allowing subsequent pathological investigations (**Fig. 1*b***).

The DeepHRD breast cancer FF model exhibited an overall performance with an AUC of 0.81 ([0.77-0.85] 95% Confidence Interval (CI); **Fig. 2*a***) on the held-out TCGA samples. The generalizability of the FF model was externally validated by applying it to 116 primary breast cancer slides from the Clinical Proteomic Tumor Analysis Consortium (*37*) and 419 primary breast tumors from the Molecular Taxonomy of Breast Cancer International Consortium (*38*) (**fig. S2*a***) resulting in an AUC of 0.76 ([0.71-0.82] 95% CI; **Fig. 2*a***). Notably, while HRD is enriched in luminal B, basal-like, and Her2 enriched breast cancers (**fig. S1*a***), DeepHRD was able to distinguish HR deficiency and proficiency across all subtypes (**Fig. 2*b***). The DeepHRD breast cancer FFPE model exhibited an AUC of 0.81 ([0.77-0.86] 95% CI; **Fig. 2*c***) on the held-out TCGA samples, which was identical to the flash frozen model. These results indicate that the fixation procedure and differences in staining coloration have minimal effects on the performance of predicting HRD status directly from breast cancer tissue slides.

**Fig. 2.**
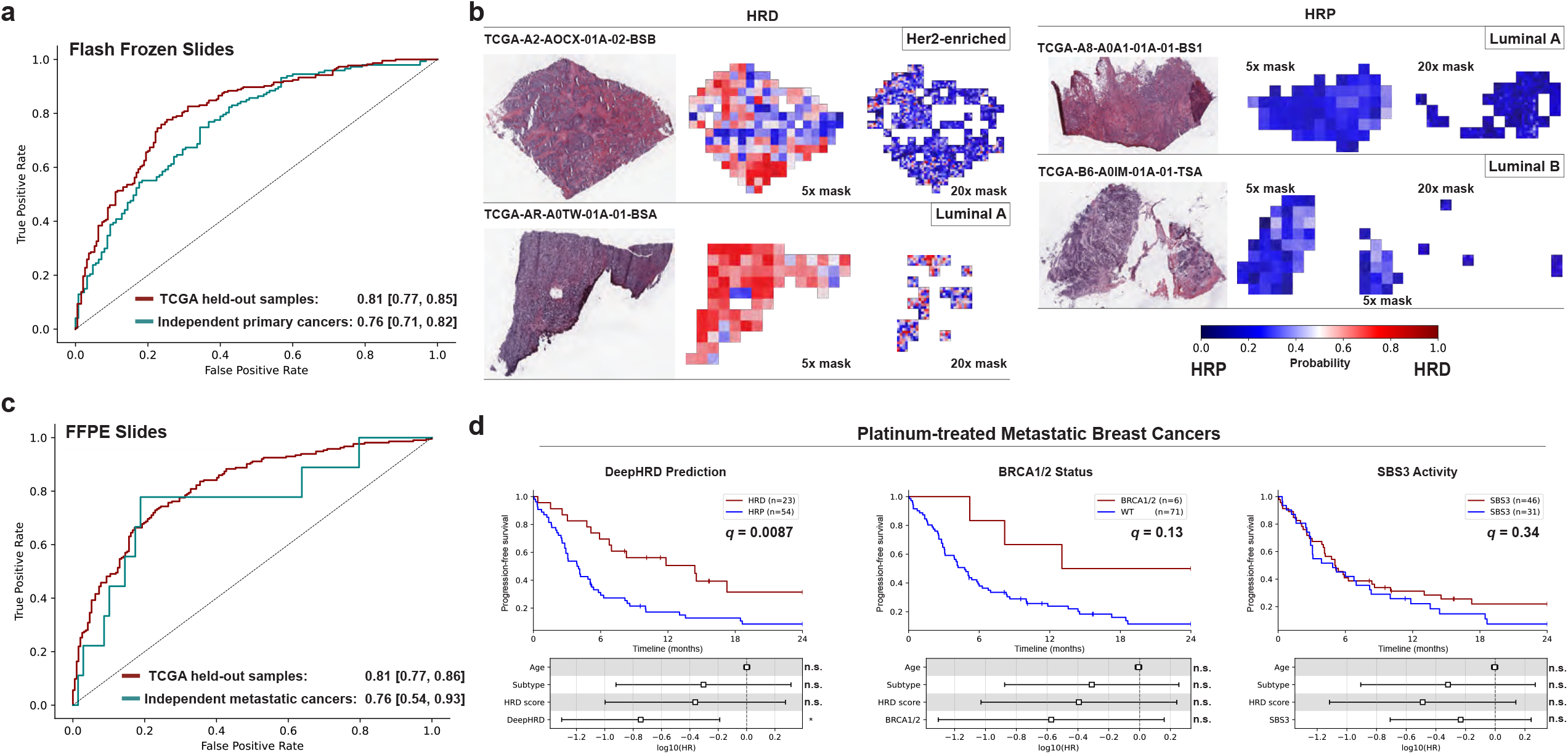
DeepHRD for detecting homologous recombination deficiency and predicting response to treatment in primary and metastatic breast cancer. ***a*)** The receiver operating characteristic curves (ROCs) for classifying homologous recombination deficiency (HRD) in the TCGA held-out set and the independent set of primary breast cancers, encompassing the independent CPTAC and METABRIC primary breast cancer cohorts. ***b)*** Representative TCGA tissue slides are shown for both HRD and homologous recombination proficient (HRP) samples across multiple breast cancer subtypes along with the resulting predictions for each segmented tile at 5x and 20x resolutions. ***c)*** ROCs for formalin-fixed paraffin-embedded (FFPE) diagnostic model in the TCGA held-out set and for classifying metastatic breast cancer (MBC) patients who are complete responders to platinum therapy. ***d***) Kaplan-Meier survival curves for MBC patients treated with platinum chemotherapy separated by DeepHRD model predictions (*left*), *BRCA1/2* mutation status (*middle*), and SBS3 activity as detected by SigMA (*right*). Q-values are corrected after considering breast cancer subtype, age at diagnosis, and the standard-of-care binary HRD classification score ≥42 (*i*.*e*., HRD score). Cox regression showing the log_10_-transformed hazard ratios are shown with their 95% confidence intervals (*bottom*). Q-values less than or equal to 0.05 are annotated with * while q-values above 0.05 are annotated with n.s. (*i*.*e*., non-significant).

Importantly, the FFPE model was capable of distinguishing metastatic breast cancers (MBCs), part of an independent clinical cohort, that had a complete response to platinum chemotherapy (*n*=9) from MBCs having only a partial or no response to treatment (*n*=68) with an AUC of 0.76 ([0.54-0.93] 95% CI; **Fig. 2*c***; **fig. S2*b***). Additionally, clinical response to platinum-based therapy and progression-free survival were assessed using the Response Evaluation Criteria in Solid Tumors, version 1.1 (RECIST 1.1; **fig. S2*b***) (*36*). Separating the MBCs treated with platinum based upon DeepHRD’s prediction revealed a median progression-free survival of 14.4 months for HRD patients and 3.9 months for HRP patients (p-value=0.0019, log-rank test). The model’s predictive value was consistent after correcting for breast cancer subtype, age of diagnosis, and the genomic HRD score with a hazard ratio of 0.47 ([0.27-0.83] 95% CI; q-value=0.0087; **Fig. 2*d***). Further, DeepHRD captured 7 of the 9 complete responders to platinum treatment. In comparison, neither the separation based upon *BRCA1/2* mutations nor detecting the HRD-associated signature SBS3 resulted in a significant difference in progression-free survival (q-value=0.13 and q=0.34, respectively; **Fig. 2*d***). While the small sample size of *BRCA1/2* mutated tumors (∼8% of MBCs) influenced the significance levels compared to wild-type tumors, the predictions from DeepHRD captured 4-fold more platinum sensitive samples. Lastly, the tissue slides from the MBC were digitalized using a Hamamatsu Photonics Nanozoomer system, while all other cohorts were digitalized using an Aperios ScanScope system, further demonstrating the generalizability of DeepHRD.

Ovarian cancer patients have traditionally received first-line platinum chemotherapies making them ideal to evaluate whether HRD predictions from tissue slides may have a direct clinical benefit. To test whether DeepHRD can be used for other cancer types, we trained an independent FF ovarian cancer model by performing transfer learning on the TCGA ovarian cancer cohort (*n*=589) using the pretrained weights and biases generated from the FF breast cancer model with the convolutional weights and biases frozen during training (**Fig. 3*a***; **fig. S1**). A similar training approach employed for the breast cancer models was utilized for the ovarian cancer model (**Supplementary Materials**). To assess the ability of the DeepHRD ovarian model to separate individuals benefiting from treatment with platinum chemotherapy, the model was applied to a held-out set of 66 high-grade serous ovarian cancers that received treatment with first-line platinum chemotherapy. Patients predicted to be HRD had a median survival of 4.6 years, while those predicted to be HRP had a median survival of 3.2 years with a hazard ratio of 0.45 ([0.22-0.90] 95% CI; q-value=0.024) after correcting for the stage of the cancer, age, and the genomic HRD score (**Fig. 3*b***). In comparison, we observed a worse separation when using a base model without transfer learning (HR=0.53 [0.26-1.07] 95% CI; q-value=0.076; **Fig. 3*c***), suggesting that the transfer learning provides a benefit when attempting to train AI-based approaches on smaller datasets. Consistent with the breast cancer cohort, neither separation based on mutations in *BRCA1/2* nor detecting the HRD-associated signature SBS3 resulted in a significant difference in survival (q-values>0.10; **Fig. 3*c***).

**Fig. 3.**
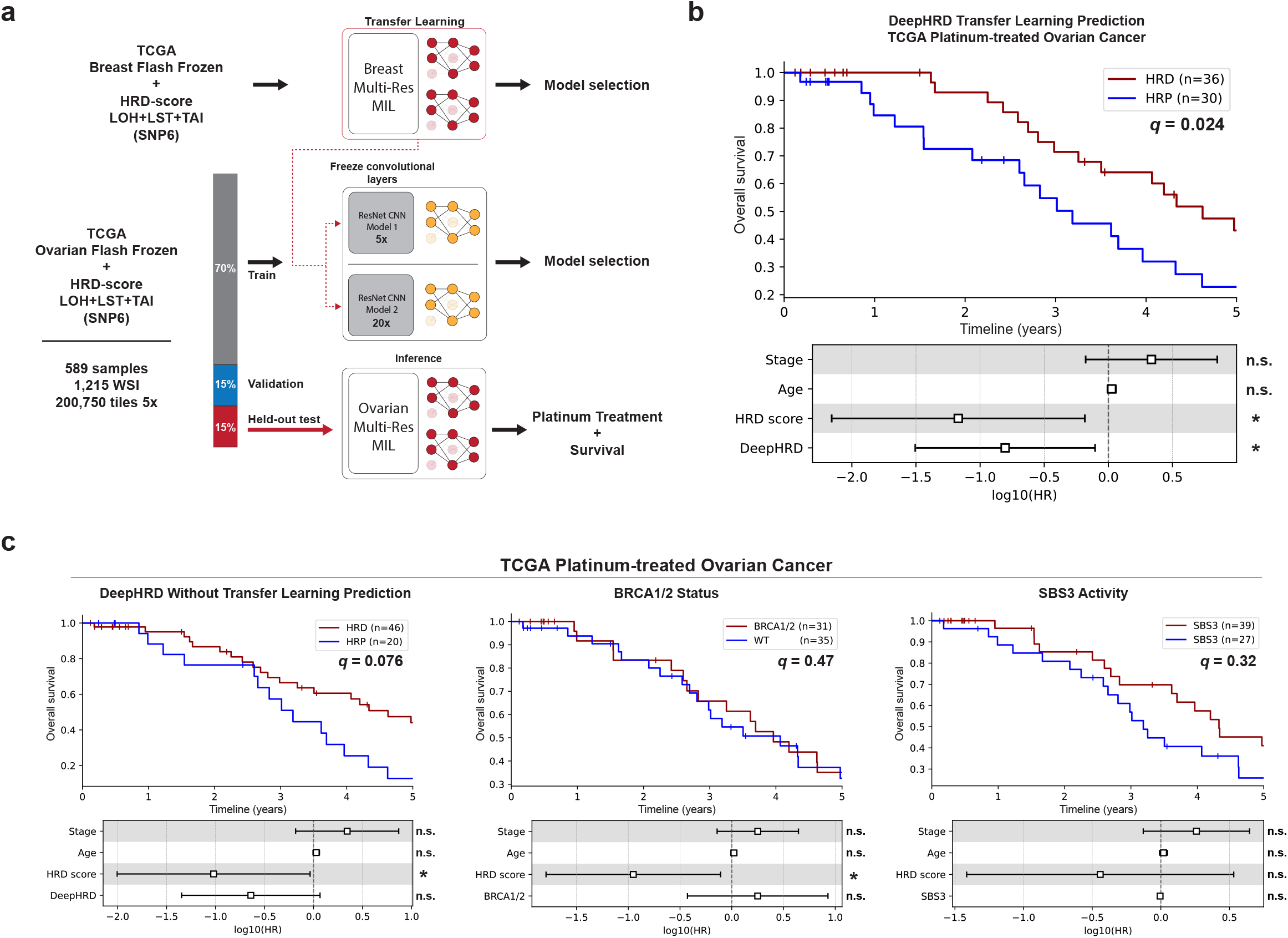
DeepHRD transfer learning in ovarian cancer for predicting response to platinum treatment. ***a)*** Schematic demonstrating the transfer learning method to train an ovarian homologous recombination deficiency (HRD) model from whole-slide H&E image (WSI) using a pretrained breast DeepHRD model. The pretrained flash-frozen breast model is used to initiate the weights and biases of all parameters in the ovarian model. HRD-scores are calculated from SNP6 genotyping microarray by deriving loss of heterozygosity (LOH), large-scale transitions (LST), and telomeric allelic imbalance (TAI). ***b)*** Kaplan-Meier survival curves comparing the outcomes of patients treated with platinum chemotherapy split by the prediction of the DeepHRD transfer learning model. ***c)*** Kaplan-Meier survival curves comparing the outcomes of platinum-treated patients split by the base model predictions with no transfer learning applied (*left*), *BRCA1/2* mutation status (*middle*), and SBS3 activity as detected by SigMA (*right*). Q-values are corrected after considering ovarian cancer stage, age at diagnosis, and the standard-of-care binary HRD classification score ≥63 (*i*.*e*., HRD score). Cox regression showing the log10-transformed hazard ratios are shown with their 95% confidence intervals (*bottom*). Q-values less than or equal 0.05 are annotated with * while q-values above 0.05 are annotated with n.s. (*i*.*e*., non-significant).

## DISCUSSION

The development of DeepHRD prediction models for breast and ovarian cancers demonstrates the practicality of deploying AI-based guidance into clinical diagnostics and precision medicine workflows. Results across multiple external cohorts indicate that the platform is applicable to routinely sampled tissue blocks and generalizable across different cancers, digital scanning systems, and tissue fixation procedures. DeepHRD’s performance was consistent across primary and metastatic breast cancers and, by incorporating transfer learning, the model was also applicable to serous ovarian cancer. Since HRD is a complementary biomarker guiding the use of platinum therapies and an FDA-approved companion diagnostic for the use of PARP inhibitors (*15-17*), the performance of our DeepHRD platform has direct implications for the treatment of other cancer types with known HR-deficiencies (*19*), notably within pancreatic adenocarcinomas. Despite clear benefit from first-line platinum therapy in HRD-positive patients with this refractory disease, there is a 3 to 6 week turn-around for genomic testing which is not appropriate for an advanced pancreatic cancer with a median progression-free survival of 6 months (*25*). Further, access to HRD genomic testing is even more limited in developing countries (*15*). With increasing clinical evidence for the treatment of HRD tumors with platinum salts and PARP inhibitors and the limitations of existing genomic tests, there is a need to develop novel frameworks to guide the current standard-of-care for HRD tumors (*23*).

Recently, deep learning AI approaches have demonstrated the ability to detect genomic alterations directly from H&E images, including biomarkers related to patient outcome, which could be leveraged for pre-screening tests. While demonstrating a high-concordance in predicting genomic HRD, DeepHRD was also capable of directly predicting individual patient outcome to HRD-targeted therapy using response and progression-free survival based on RECIST criteria. Furthermore, DeepHRD provided better prediction of clinical response and progression-free survival to platinum therapies than existing genomic biomarkers. Notably, our approach captured patients with *BRCA1/2* wild-type tumors who responded to platinum therapy, thus, identifying 4-fold more responders than *BRCA1/2* mutation-testing alone (**Fig. 2*d***). These results demonstrate that molecular assays, traditionally used for assessing HRD in a clinical setting, can be substituted and/or complemented with AI-based deep learning models for predicting clinical response from conventional diagnostic histopathological slides.

While there has been a recent explosion of deep learning methods applied to digital pathology (*28*), the immediate translation into clinical practice has been limited by the high costs associated with acquiring infrastructure for routinely capturing digital H&E slides (*27, 39*). With the development of tertiary scanning services seeking to alleviate these overhead costs, a recent study has shown a potential alternative approach for utilizing analogous deep learning AI-models to make predictions from photographs taken directly by hand-held devices (*40*). In coordination with the optimization of lightweight deep learning architectures, using images from a smartphone attached to the ocular lens of a conventional light microscope promises inexpensive, efficient, and accurate deep-learning read-outs within seconds of preparing an H&E slide. By relying on smartphone microscopy images, this transition would provide AI-based diagnostic solutions for equitable and efficient clinical management for cancer patients across globally diverse socioeconomic groups.

## Data Availability

The collection of flash frozen and formalin-fixed paraffin-embedded (FFPE) slides from TCGA along with all clinical features were downloaded from the Genomic Data Commons (GDC; https://gdc.cancer.gov/). The collection of flash frozen slides from CPTAC were downloaded from The Cancer Imaging Archive (TCIA), and the genomics data was downloaded from the GDC. The collection of images from METABRIC and the associated SNP6 genotyping microarray data were downloaded from European Genome-Phenome Archive (EGA) with accession numbers: EGAD00010000270 and EGAD00010000266. The collection of samples from the metastatic breast cancer cohort are available on Sequence Read Archive (SRA) repository under BioProject accession number PRJNA793752.

https://ega-archive.org/datasets/EGAD00010000270

https://ega-archive.org/datasets/EGAD00010000266

https://www.ncbi.nlm.nih.gov/sra/SRX13584185

## Acknowledgements

The results shown are in part based upon data generated by the TCGA Research Network: http://cancergenome.nih.gov/. Additional data used in this publication were generated by the Clinical Proteomic Tumor Analysis Consortium (CPTAC) and the Molecular Taxonomy of Breast Cancer International Consortium (METABRIC). The computational analyses reported in this manuscript have utilized the Triton Shared Computing Cluster at the San Diego Supercomputer Center of UC San Diego.

## Author contributions

ENB and LBA designed the overall study. ENB performed all analyses with help from AA, MDG, LG, SML, SL, and LBA. Specifically, AA assisted in the calculation and interpretation of the genomically-derived HRD scores. MDG assisted in the processing of the digital images. LG and SL assisted in the analysis and interpretation of the metastatic breast cancer cohort. SML assisted in the interpretation and analysis of the survival and clinical associations. ENB and LBA wrote the manuscript with help and input from all other authors. All authors read and approved the final manuscript.

## Data and materials availability

All datasets utilized in this study were previously generated and publicly available. These data can be access through the accession codes listed in **Supplementary Materials**. The sources code for DeepHRD will be made publicly available upon the publication of this study.

