## Supplementary Materials for "Deep learning predicts HRD and platinum response from histology slides in breast and ovarian cancer"

**The PDF file includes:**

Materials and Methods
Figs. S1 to S2
References

### Materials and Methods

#### ***Data sources***

The collection of flash frozen and formalin-fixed paraffin-embedded (FFPE) slides from The Cancer Genome Atlas project (TCGA) along with all clinical features were downloaded from the Genomic Data Commons (GDC; https://gdc.cancer.gov/). The collection of flash frozen slides from Clinical Proteomic Tumor Analysis Consortium (CPTAC) were downloaded from The Cancer Imaging Archive (TCIA), and the genomics data was downloaded from the GDC. The collection of images from Molecular Taxonomy of Breast Cancer International Consortium (METABRIC) and the associated SNP6 genotyping microarray data were downloaded from EGA (accession numbers: EGAD00010000270 and EGAD00010000266). The predicted cancer subtype for a subset of the TCGA breast cancer cohort were obtained from a previous study that utilized the 50-gene PAM50 model (*1*). HRD scores for the TCGA breast and ovarian cancers were obtained from a previous study (*2*). The 77 patients from the clinical cohort of whole exome sequenced metastatic breast cancers were enrolled between June 2018 and March 2020 and all received at least one line of platinum chemotherapy. All clinical evaluations were determined locally at the Georges Francois Leclerc Cancer Center as previously reported (*3*).

#### ***Data preprocessing***

Each of the whole-slide images (WSIs) was segmented into 256x256 tiles at 5x and 20x magnifications containing 2µm per pixel and 0.5µm per pixel, respectively. Blurry tiles and those with less than 80% of pixels representing tissue were removed from all training and testing cohorts. To filter blurry tiles, a Laplacian filter was applied to each tile using a 3x3 kernel, and all tiles with a variance less than 0.02 were removed from the remaining analysis. All green, red, and blue pen marks and other annotation artifacts were removed by thresholding on the RGB color channels within each pixel.

***Calculating HRD scores***

HRD scores were calculated as previously reported using scarHRD (*2, 4*). Specifically, the HRD score is the summation of the telomeric allelic imbalance score, loss of heterozygosity score, and large-scale transitions scores calculated for each patient using ASCAT-derived (*5*) copy number calls from SNP6 genotyping microarrays. The HRD scores for the CPTAC breast cancer samples were calculated based on copy number calls derived from whole-exome sequencing using Sequenza (*6*), which has been shown to result in analogous distributions of HRD scores to HRD scores calculated using ASCAT-derived (*5*) copy number calls from SNP6 genotyping microarrays.

For TCGA primary cancers, the pathogenicity of *BRCA1/2* mutations was determined using InterVar as previously described (*7*). For metastatic breast cancer, *BRCA1/2* mutations were determined by screening variants across multiple databases as previously reported (*3*). All pathogenic variants were considered deleterious. The presence of signature SBS3 was identified based on high confidence predictions from SigMA (*8*), a machine learning tool for detecting HRD status based on SBS3. SigMA has 1% false positive rate and 50% sensitivity (*8*).

***Soft labeling of whole-slide images***

For both TCGA breast and ovarian cancer cohorts, soft labeling was applied to the HRD scores using specified thresholds for samples labeled confidently as HRD or HRP. These thresholds were determined by first splitting samples within each cancer type into HRD and HRP partitions using a single cutoff (HRD>=30 for breast and HRD>=63 for ovarian). The median values of the two resulting partitions of samples for each cancer type were used to set the range of confident HRD and HRP thresholds. All intermediate HRD scores were modeled as a probability using a quadratic function (**equation 1**).

$$HRD_{Adjust} = \begin{cases} 2 * \left(\frac{HRD_{thresh} - x}{HRD_{thresh} - HRP_{thresh}} - 0.5\right)^2 + 0.5 & \boldsymbol{if\ HRD_{median} \le x < HRD_{thresh}} \\ 1 - \left[2 * \left(\frac{HRD_{thresh} - x}{HRD_{thresh} - HRP_{thresh}} - 0.5\right)^2 + 0.5\right] & \boldsymbol{if\ HRP_{thresh} \le x < HRD_{median}} \end{cases} \quad \textbf{(1)}$$

Specifically, HRD scores above 50 were considered HR-deficient and scores below 10 were considered proficient in the breast cancer cohorts. All intermediate scores were modelled as a probability of being deficient or proficient with an equal probability of both conditions at an HRD score of 30 (**equation 2**).

$$HRD_{Adjust} = \begin{cases} 2 * \left(\frac{50 - x}{50 - 10} - 0.5\right)^2 + 0.5 & \boldsymbol{if\ 30 \le x < 50\ (breast)} \\ 1 - \left[2 * \left(\frac{50 - x}{50 - 10} - 0.5\right)^2 + 0.5\right] & \boldsymbol{if\ 10 \le x < 30\ (breast)} \end{cases} \quad \textbf{(2)}$$

Within the TCGA ovarian cohort, HRD scores above 73 were considered deficient and scores below 53 were considered proficient with the intermediate probabilities centered at 63 (**equation 3**).

$$HRD_{Adjust} = \begin{cases} 2 * \left(\frac{73-x}{73-53} - 0.5\right)^2 + 0.5 & \boldsymbol{if\ 63 \le x < 73\ (ovarian)} \\ 1 - \left[2 * \left(\frac{73-x}{73-53} - 0.5\right)^2 + 0.5\right] & \boldsymbol{if\ 53 \le x < 63\ (ovarian)} \end{cases} \quad \textbf{(3)}$$

***Model training and testing***

Prior to training, the number of HRD and HRP samples were balanced in all breast cancer subtypes using the PAM50 model classifications to normalize for specific breast cancer subtypes being enriched or depleted of HRD samples (*1*). All samples without annotated PAM50 subtype labels were considered as missing and were also balanced for the number of HRD and HRP cases (**fig. S1*a***). Soft labelling was incorporated to prevent overfitting during training and to account for ambiguity in the ability of the HRD score to classify true HRD samples. The entirety of training and testing was performed using the machine learning Python framework Pytorch (v.1.5.0). For both resolution models, the Adam optimizer was used for training with a learning rate of $10^{-3}$, a weight decay of $10^{-4}$, and minibatches consisting of 64 tiles. Each model was initiated using the ResNet18 (*9*) architecture that was pretrained on the ImageNet (http://www.image-net.org/) database and was trained for a maximum of 200 epochs. All convolutional weights were frozen during training. Dropout within the fully connected layers and early stoppage during training were incorporated to prevent overfitting. Individual DeepHRD prediction thresholds for each trained model were selected based upon the classification performance on the held-out test sets.

After training the 5x resolution models, a final inference pass is performed on all slides. All features from a single WSI were selected from the penultimate layer of the feature extractor and projected into a lower dimensional latent space using principal component analysis. K-means clustering was used to automatically select regions of interests (ROIs) for retiling at 20x magnification. The number of clusters was determined by selecting the solution with the maximum silhouette coefficient. The cluster containing the tile with the highest prediction probability was used to select the ROIs. All tiles belonging to this cluster, and which had a silhouette score greater than the 95% quantile of all silhouette scores for the given WSI were chosen as the final ROIs. Each ROI was then tiled into 256x256 pixel sub-tiles at 20x magnification. This results in 16 tiles at 20x magnification for each ROI at a 5x magnification. The top 25 tiles were averaged to calculate a final prediction score at a given resolution during an inference pass of a WSI. To perform an inference pass of the model, a single WSI image is processed across 10 iterations with a random dropout probability of 0.20 for all nodes within the fully connected layers.

***Transfer learning***

The weights collected from the final models trained to detect HRD from flash frozen breast slides were used to initiate the model weights for the ovarian model known as transfer learning. All other training procedures were consistent with the training of the breast cancer models. The held-out internal validation set was used to perform survival analysis based upon prior treatment with platinum chemotherapy. There were not enough FFPE slides for the ovarian cohort for training and testing a DeepHRD model for FFPE ovarian cancer samples.

***Visualizing DeepHRD predictions***

Once successfully trained, DeepHRD is used to make predictions for individual whole-slide images. When performing the multi-resolution inference, DeepHRD generates HRD probabilities for each tile at 5x magnification and for each tile within the automatically selected regions of interest at 20x magnification. Using the location of the original tiles, the probabilities can be mapped back to the original location within the whole-slide image to visualize the regional patterns that are influencing the final model prediction.

***Survival analysis***

Survival analysis was performed using the Lifelines Python package (v.0.24.4.). For both the metastatic breast cancer (MBC) and the TCGA ovarian cohorts, samples were partitioned based upon the prediction from each respective DeepHRD model. Only samples that were treated with platinum chemotherapy were considered in the survival comparisons. Survival curves were compared using a log-rank test. Hazard ratios were calculated from Cox regressions (*10*) after correcting for age of diagnosis, primary breast cancer subtype, and genomic HRD score within the MBC cohort and age of diagnosis, ovarian cancer stage, and genomic HRD score within the TCGA ovarian cohort. Median survival was calculated as the time at which the chance of surviving beyond that point is 50%. The Benjamini Hochberg's procedure was used for multiple hypothesis testing corrections (*11*).

***Statistical analysis***

DeepHRD's performance was evaluated by calculating the area under the receiving operating curve (AUC). Confidence intervals were derived using non-parametric resampling. Comparisons across survival curves were implemented using a log-rank test. Multivariate analyses were performed to calculate hazard ratios using Cox regressions. The Benjamini Hochberg's procedure was used for multiple hypothesis testing corrections. All performance metrics were calculated using the scikit-learn Python package (v.0.22.1). Confidence intervals were calculated using non-parametric resampling. Standard error bars were calculated using the NumPy Python package (v.1.18.1).

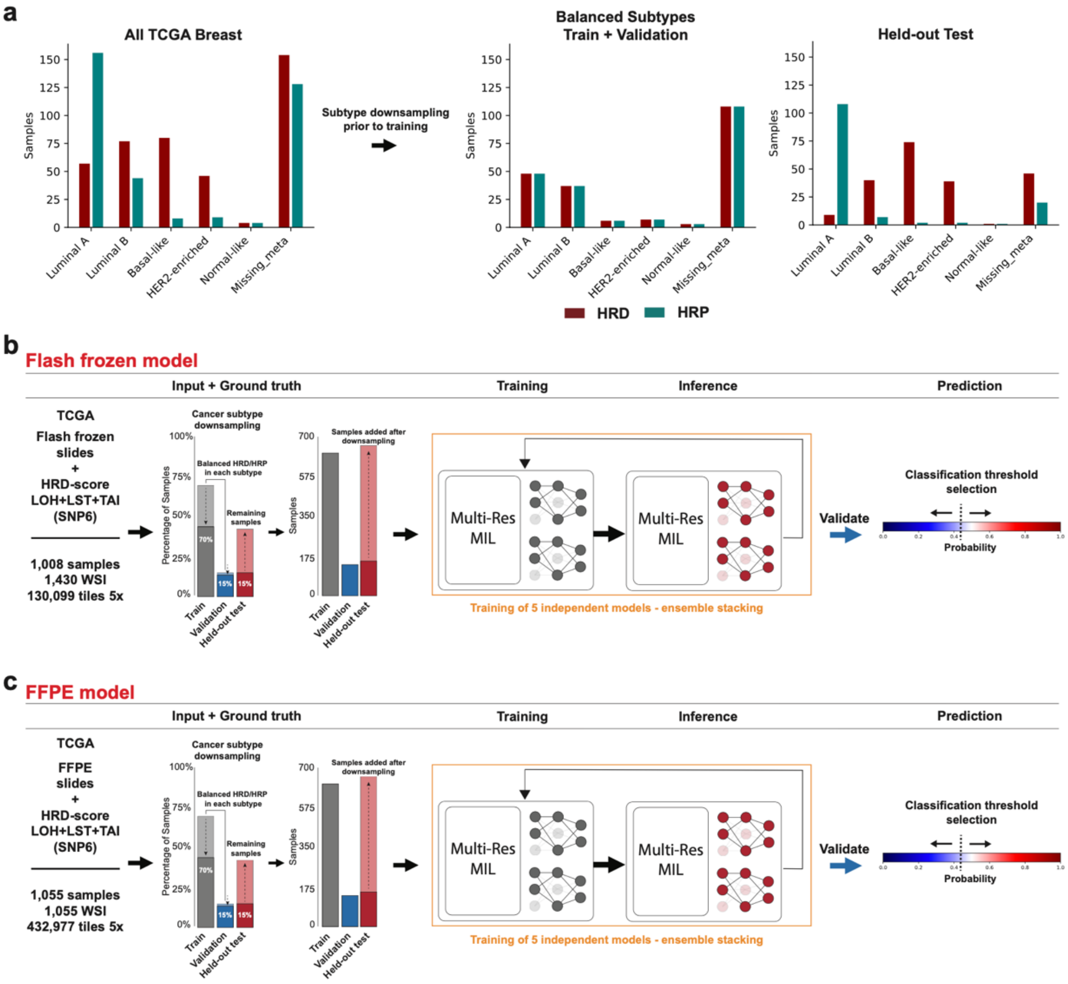


**Fig. S1. Workflow for training DeepHRD models independently for digitalized flash frozen and FFPE breast cancer slides.** ***a***) Prior to training, the number of HRD and HRP samples within each breast cancer subtype were balanced using all available PAM50 annotations (*1*). The collection of flash frozen ***b)*** and formalin-fixed paraffin-embedded (FFPE) ***c)*** slides for the TCGA breast cancer cohort were used to train two independent DeepHRD models. Prior to training, the number of HRD and HRP samples were balanced within each breast cancer subtype. All downsampled individuals were added to the internal held-out test set. The validation sets were used to optimize the classification thresholds.

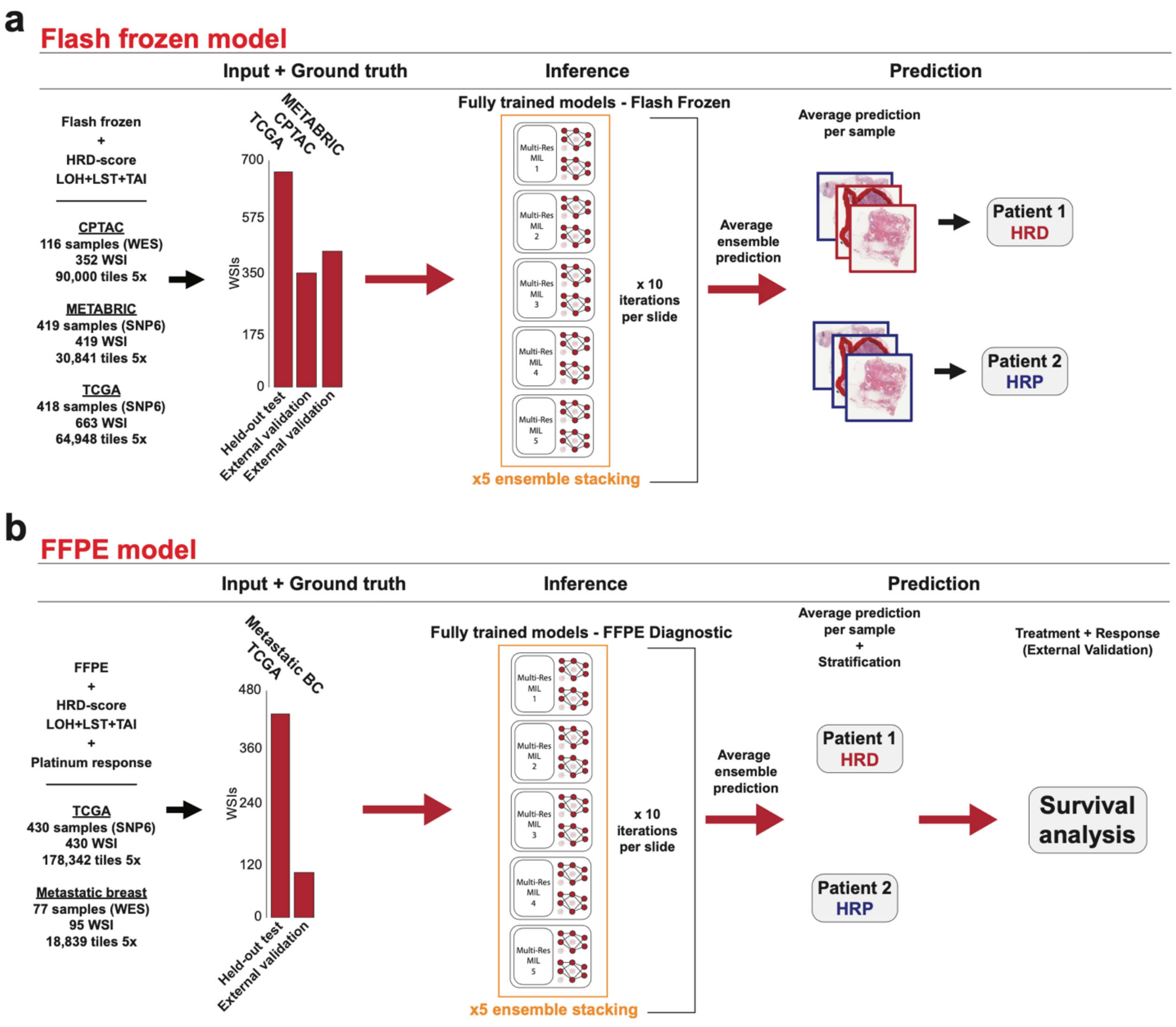


**Fig. S2. Workflow for testing the performance of DeepHRD models for digitalized flash frozen and FFPE breast cancer slides.** ***a)*** The collection of breast cancers from CPTAC and METABRIC were used to independently validate the flash frozen breast cancer model. The DeepHRD prediction scores were averaged for samples with multiple images. ***b)*** An independent collection of metastatic breast cancers treated with platinum chemotherapy was used to validate the formalin-fixed paraffin-embedded (FFPE)breast cancer model based upon individual patient response to therapy.
